# Development and validation of a questionnaire to measure attitudes toward COVID-19 vaccination and pandemic

**DOI:** 10.1101/2021.09.30.21264344

**Authors:** Petros Galanis, Irene Vraka, Olga Siskou, Olympia Konstantakopoulou, Aglaia Katsiroumpa, Ioannis Moisoglou, Daphne Kaitelidou

## Abstract

**Background:** Accurate measurement of individuals’ attitudes toward COVID-19 vaccination and pandemic is critical to understand the way that people respond during a major crisis such as the COVID-19 pandemic.

**Objective:** To develop and validate a questionnaire to assess attitudes toward COVID-19 vaccination and pandemic.

**Methods:** We performed a reliability and validity study in a sample of the general population in Greece. Data were collected online through social media between 15 August and 7 September 2021. Thus, a convenience sample was obtained. Reliability and validity of the questionnaire were assessed with a Delphi study, an exploratory factor analysis, and a test-retest study. Also, we calculated Cronbach’s coefficient alpha for the factors that emerged from the exploratory factor analysis.

**Results:** The final study included 1959 adults from the general population in Greece. Our four-factor model explained 73% of the variance and confirmed out initial hypothesis regarding the factors of the questionnaire. In particular, we found the following four factors: (a) fear against the COVID-19 (five items), (b) information regarding the COVID-19 (two items), (c) compliance with hygiene measures (two items), and (d) trust in COVID-19 vaccination (seven items). Cronbach’s coefficients alpha for the four factors that emerged from the exploratory factor analysis were greater than 0.82. Pearson’s correlation coefficients for the 16 items and the four factors were greater than 0.67 (p-value<0.001 in all cases).

**Conclusions:** We developed a reliable and valid questionnaire to measure attitudes toward COVID-19 vaccination and pandemic. Further studies should be conducted to expand our knowledge and infer more valid results.

## Introduction

The personal, social and economic impact of the Coronavirus disease 2019 (COVID-19) pandemic is enormous. Until October 2021, the COVID-19 pandemic has caused more than 4.8 million deaths and 235 million cases (Worldometer, 2021). Moreover, the COVID-19 pandemic has an adverse effect on the mental health of the general population causing depression, anxiety, post-traumatic stress disorder, sleep disturbances, psychological distress, etc. (Ayubi et al., 2021; Hossain et al., 2021; Pashazadeh Kan et al., 2021; Zhang et al., 2021). Additionally, adverse psychological and physical symptoms due to the COVID-19 pandemic are more frequent among high-risk groups such as healthcare professionals, patients, students and teachers (Galanis, Vraka, Fragkou, et al., 2021a, 2021b; Y Li et al., 2021; Mahmud et al., 2021; Ozamiz-Etxebarria et al., 2021; Sterina et al., 2021; Wang et al., 2021).

From early 2021, the ability to vaccinate against the COVID-19 with safe and effective vaccines is the best hope to control the pandemic (Baden et al., 2021; Haas et al., 2021; Polack et al., 2020). Unfortunately the intention to vaccinate against the COVID-19 is low not only in the general population but also among healthcare professionals with some countries showing higher hesitancy rates compared with others (Cascini et al., 2021; Galanis et al., 2020; Galanis, Vraka, Siskou, et al., 2021a, 2021b; Luo et al., 2021; Salomoni et al., 2021; Snehota et al., 2021). Moreover, several factors influence the intention to vaccinate against the COVID-19 such as socio-demographic characteristics (e.g. gender, age, race, and educational level), morbidity, attitude towards a COVID-19 vaccine and vaccination in general, fear and self-perceived risk about COVID-19, etc.

Until now, several questionnaires and scales have created to examine psychological impact of COVID-19 and mainly examine stress, anxiety or fear (Ahorsu et al., 2020; Arpaci et al., 2020; Feng et al., 2020; Lee, 2020; Lins and Aquino, 2020; Repišti et al., 2020; Taylor et al., 2020; Voitsidis et al., 2021). Also, several other questionnaires have developed to measure knowledge, attitudes, and practices about the COVID-19 (Bekele et al., 2020; Dardas et al., 2020; Patelarou et al., 2020; Saadatjoo et al., 2021; Srichan et al., 2020). To the best of our knowledge, no fully validated questionnaire to measure attitudes toward COVID-19 vaccination and pandemic exists. Thus, we aimed to develop and validate a questionnaire to assess attitudes toward COVID-19 vaccination and pandemic.

## Methods

### Development of the questionnaire

First, a systematic literature review was conducted on the attitudes of individuals toward COVID-19 vaccination and pandemic, resulting in a pool of relevant items. The themes that emerged were as follows: (a) fear against the COVID-19, (b) information regarding the COVID-19, (c) compliance with hygiene measures, and (d) trust in COVID-19 vaccination. A Delphi study was then conducted involving eight experts; two physicians, two epidemiologists, two nurses and two psychologists (Galanis, 2018). In this case, the content validity ratio was calculated for the initial pool of items and the items with a content validity ratio greater than 0.85 were included in the questionnaire distributed to the participants (Galanis, 2013). The final questionnaire comprised 16 items. Responses in items ranged from 0 = “totally disagree” to 10 = “totally agree”.

### Pilot study

A pilot study was conducted with 30 participants aged 18 to 75 years in which face validity was tested. Specifically, participants were asked to answer the 16 items of the questionnaire and to report possible errors, omissions, ambiguities and misinterpretations. Minimal verbal corrections were made, resulting in the final version of the questionnaire.

### Final study

The final study included 1959 adults from the general population in Greece. Data were collected online through social media between 15 August and 7 September 2021. Thus, a convenience sample was obtained. Participants were informed about the purpose and methodology of the study and consented to participate. Data was collected anonymously and no personal data was collected. Institutional ethical approval from the Department of Nursing, National and Kapodistrian University of Athens (reference number; 370, 02-09-2021) was obtained prior to conducting the study.

### Statistical analysis

We used exploratory factor analysis to assess the construct validity of the questionnaire and to find out possible underlying factors. In particular, we used the varimax rotation method to assess correlation between the 16 items. In that case, acceptable level for factor loading was set at 0.4, while for eigenvalues was set at 1. We calculated the Kaiser-Meyer-Olkin measure to assess the adequacy of the sample size with a value >0.8 considered to be acceptable. Moreover, we performed Bartlett’s test of sphericity and a p-value <0.05 indicates that the correlation matrix is appropriate for exploratory factor analysis.

We assessed reliability of the questionnaire in two ways. First, we performed a test-retest study with 50 participants that filled the questionnaire two times with a time interval of 10 days. Then we calculated, Pearson’s correlation coefficient for the 16 items with values >0.6 considered to be satisfactory. Moreover, we calculated Cronbach’s coefficient alpha for the factors that emerged from the exploratory factor analysis with values greater than 0.7 indicate an acceptable level of internal reliability.

We used numbers (percentages) to present categorical variables and mean (standard deviation) to present continuous variables. All tests of statistical significance were two-tailed, and p-values < 0.05 were considered as statistically significant. Statistical analysis was performed with the IBM SPSS 21.0 (IBM Corp. Released 2012. IBM SPSS Statistics for Windows, Version 21.0 Armonk, NY, USA).

## Results

### SDemographic characteristics

Detailed demographic profile of the participants is shown in Table 1. Mean age of the participants was 41.5 years old, while most of them were females (75.3%) and married (64.3%). Among the participants, 81% had a University degree. Nine point five percent of the participants were diagnosed with COVID-19. During the COVID-19 pandemic, 30.9% of the participants lived with elderly people or vulnerable groups during the COVID-19 pandemic.

**Table 1.** Demographic characteristics of the participants.

| **Characteristics** | **N** | **%** |
| --- | --- | --- |
| Gender |  |  |
| Females | 1476 | 75.3 |
| Males | 483 | 24.7 |
| Age (years)<sup>a</sup> | 41.5 | 10.6 |
| Marital status |  |  |
| Singles | 553 | 28.2 |
| Married | 1260 | 64.3 |
| Widowed | 135 | 6.9 |
| Divorced | 11 | 0.6 |
| Educational level |  |  |
| Elementary school | 28 | 1.2 |
| High school | 345 | 17.6 |
| University degree | 1586 | 81.0 |
| Previous COVID-19 diagnosis |  |  |
| No | 1773 | 90.5 |
| Yes | 186 | 9.5 |
| Living with elderly people or vulnerable groups during the COVID-19 pandemic |  |  |
| No | 1354 | 69.1 |
| Yes | 605 | 30.9 |

<sup>a</sup> mean, standard deviation

### Factor analysis

The p-value for Bartlett’s test of sphericity was <0.001, while the Kaiser–Meyer–Olkin measure was 0.88. Thus, our sample was adequate to perform the exploratory factor analysis. Table 2 presents the detailed results of the exploratory factor analysis. We found four factors including all the 16 items of our questionnaire. Our four-factor model explained 73% of the variance and confirmed out initial hypothesis regarding the factors of the questionnaire. In particular, we found the following four factors: (a) fear against the COVID-19 (five items), (b) information regarding the COVID-19 (two items), (c) compliance with hygiene measures (two items), and (d) trust in COVID-19 vaccination (seven items). The only item with a reverse scoring was the “I am worried about the side effects that COVID-19 vaccines can have”.

**Table 2.** Exploratory factor analysis for the 16 items of the study questionnaire.

| Item | Factors |  |  |  |
| --- | --- | --- | --- | --- |
|  | 1 | 2 | 3 | 4 |
|  | Fear against the COVID-19 | Information regarding the COVID-19 | Compliance with hygiene measures | Trust in COVID-19 vaccination |
| I am afraid that I will be infected with the coronavirus | 0.86 |  |  |  |
| I am afraid that my family and friends will be infected with the coronavirus | 0.86 |  |  |  |
| I am afraid that I will get seriously ill from the COVID-19 | 0.82 |  |  |  |
| I am afraid that my family and friends will get seriously ill from the COVID-19 | 0.86 |  |  |  |
| I believe that the COVID-19 is an extremely severe disease | 0.43 |  |  |  |
| I am aware of the coronavirus and the COVID-19 |  | 0.87 |  |  |
| I am aware of the COVID-19 vaccines |  | 0.88 |  |  |
| I follow personal hygiene measures for the coronavirus (e.g. hand washing) |  |  | 0.87 |  |
| I follow public health measures for the coronavirus (e.g. use of mask, social distancing) |  |  | 0.89 |  |
| I trust government officials regarding the information they provide on COVID-19 vaccines |  |  |  | 0.72 |
| I trust experts regarding the information they provide on COVID-19 vaccines |  |  |  | 0.85 |
| I trust my family physician regarding the information (s)he provides on COVID-19 vaccines |  |  |  | 0.65 |
| I am worried about the side effects that COVID-19 vaccines can have |  |  |  | 0.64 |
| I have confidence in the COVID-19 vaccines |  |  |  | 0.90 |
| I believe that COVID-19 vaccination could help control the pandemic |  |  |  | 0.89 |
| I believe that COVID-19 vaccination promotes public health |  |  |  | 0.89 |

Values express loadings.

### Reliability analysis

Cronbach’s coefficients alpha for the four factors that emerged from the exploratory factor analysis were greater than 0.82 indicating a great level of internal reliability. In particular, Cronbach’s coefficient alpha for the factor “fear against the COVID-19” was 0.87, for the factor “information regarding the COVID-19” was 0.82, for the factor “compliance with hygiene measures” was 0.86, and for the factor “trust in COVID-19 vaccination” was 0.82. Moreover, according to the Pearson’s correlation coefficient, the questionnaire showed very good reliability. Specifically, Pearson’s correlation coefficients for the 16 items and the four factors were greater than 0.67 (p-value<0.001 in all cases).

### Descriptive statistics

We calculated mean scores for each factor. In that case, we added the answers in the items of each factor and then we divided the sum with the total number of items. Thus, each factor had a total score from 0 to 10 with higher values indicate a higher level of agreement. Table 3 presents descriptive statistics for the four factors of the study questionnaire. Compliance with hygiene measures was very high, information regarding the COVID-19 was high, trust in COVID-19 vaccination was moderate to high, and fear against the COVID-19 was moderate.

**Table 3.** Descriptive statistics for the four factors of the study questionnaire

| **Factor** | **Mean** | **Standard deviation** | **Median** | **Minimum value** | **Maximum value** |
| --- | --- | --- | --- | --- | --- |
| Fear against the COVID-19 | 6.4 | 2.1 | 6.6 | 0 | 10 |
| Information regarding the COVID-19 | 8.7 | 1.4 | 9.0 | 0 | 10 |
| Compliance with hygiene measures | 7.4 | 2.0 | 8.0 | 0.1 | 10 |
| Trust in COVID-19 vaccination | 9.4 | 1.1 | 10 | 0 | 10 |

## Discussion

We performed a reliability and validity study in a sample of the general population in Greece to develop a questionnaire that measures attitudes toward COVID-19 vaccination and pandemic. Our questionnaire has proven to be reliable and valid and consists of the following four factors: (a) fear against the COVID-19, (b) information regarding the COVID-19, (c) compliance with hygiene measures, and (d) trust in COVID-19 vaccination. The questionnaire includes 16 items which were simple and understandable to the participants. For this reason, filling out the questionnaire is a straightforward process and requires only a short time (about 5 minutes).

Several questionnaires have been developed to measure knowledge, attitudes, and practices about the COVID-19 (Bekele et al., 2020; Dardas et al., 2020; Patelarou et al., 2020; Saadatjoo et al., 2021; Srichan et al., 2020). Each questionnaire has pros and cons. These questionnaires have been developed over the last 20 months and each one has attempted to capture attitudes during the period of the pandemic in which it developed. Therefore, each questionnaire has a different structure, items and factors so that it could meet the objectives set by its creators. In addition, each questionnaire has advantages and disadvantages and was created for a specific population. For this reason, it is necessary to create new questionnaires as the pandemic evolves so that the research can be better adapted to the new evidence. In this context, we created our questionnaire to measure attitudes toward COVID-19 vaccination and pandemic.

One of the four factors that we identified through exploratory factor analysis was trust in COVID-19 vaccination. Individuals’ trust in COVID-19 vaccination is critical since it could reduce vaccine hesitancy (Dubé et al., 2013; Gust et al., 2005). Vaccine hesitancy is a complicated issue and probably the main obstacle for the population to accept COVID-19 vaccines (Jaca et al., 2021; Wiysonge et al., 2021). Thus, there is a need for appropriate interventions such as transparent and reasonable COVID-19 vaccine educational campaigns and behavioral-change interventions (Lin et al., 2020; Schaffer DeRoo et al., 2020). In this context, policy makers, scientists and government officials could diminish people’s concerns for COVID-19 vaccine safety and efficacy.

Additionally, information regarding the COVID-19 was another factor that emerged from our analysis. People’s ability to detect fake news and recognize misinformation is crucial to their intention to take a COVID-19 vaccine (Montagni et al., 2021). For instance, people that not rely on social media to get information about the COVID-19 pandemic are those with a higher COVID-19 vaccine uptake rate (Barry et al., 2021). In general, COVID-19 information from most websites is a poor source of information resulting on misinformation and confusing messages (Cuan-Baltazar et al., 2020; Fan et al., 2020; Joshi et al., 2020). The COVID-19 infodemic that is created during the pandemic is major public health issue. Especially, the information regarding the COVID-19 vaccines should be treated with thoughtfulness and critical thinking.

Fear against the COVID-19 was the third factor in our four-factor model of attitudes toward COVID-19 vaccination and pandemic. Fear against the COVID-19 is associated with a higher acceptance rate of a COVID-19 vaccine (M Li et al., 2021). During the COVID-19 pandemic individuals experience extreme levels of fear of COVID-19 that resulting on psychological and physical problems, e.g. insomnia, burnout, depression, stress disorders, anxiety disorders, etc. (Cabarkapa et al., 2020; Garfin et al., 2020; Gorini et al., 2020; Nazar et al., 2020; Saracoglu et al., 2020; Shigemura et al., 2020; Sloan et al., 2020).

The final factor that we identified through the factors analysis was compliance with hygiene measures. Research has already shown that the COVID-19 pandemic affects in a positive way the personal hygiene behaviors such as hand washing (Dwipayanti et al., 2021; Hu et al., 2021; Huang et al., 2021). In general, people have modified their attitudes to confront the risk propensity of the COVID-19 pandemic. Specifically, individuals have increased their hand hygiene practices during the COVID-19 pandemic.

### Limitations

Our study had several limitations. First, our questionnaire has proven to be reliable and valid but it should be used in other populations to test further its psychometric properties since different populations may have different attitudes and cultural context. Further studies could confirm our four-factor model or could expand our findings. Second, we used a large sample of the general population in Greece but our sample was a convenience sample. Thus, further studies with more representative samples should be conducted to infer more valid results. Third, as always, creation of a questionnaire could not be exhausted in the first place. For instance, further items could be added in our questionnaire and different factors could be established.

## Conclusions

In conclusion, we developed a reliable and valid questionnaire to measure attitudes toward COVID-19 vaccination and pandemic. We came up with a four-factors model; (a) fear against the COVID-19, (b) information regarding the COVID-19, (c) compliance with hygiene measures, and (d) trust in COVID-19 vaccination. Accurate measurement of individuals’ attitudes toward COVID-19 vaccination and pandemic is critical to understand the way that people respond during a major crisis such as the COVID-19 pandemic. Thus, further studies should be conducted to expand our knowledge and infer more valid results.

## Data Availability

Data will be available after a reasonable request.

